# Differential durability of humoral and T cell immunity after two and three BNT162b2 vaccinations in adults aged >80 years

**DOI:** 10.1101/2022.02.10.22270733

**Authors:** Addi J. Romero-Olmedo, Axel Ronald Schulz, Svenja Hochstätter, Dennis Das Gupta, Heike Hirseland, Daniel Staudenraus, Bärbel Camara, Kirsten Volland, Véronique Hefter, Siddhesh Sapre, Verena Krähling, Helena Müller-Kräuter, Henrik E. Mei, Christian Keller, Michael Lohoff

**Affiliations:** Institute of Medical Microbiology and Hospital Hygiene, Philipps-University Marburg, Marburg, Germany; Deutsches Rheumaforschungszentrum Berlin, a Leibniz Institute, Berlin, Germany; Institute of Virology, Philipps-University, Marburg, Germany

## Abstract

A third mRNA-based “booster” vaccination is the favored strategy to maintain protection against SARS-CoV-2 infection. Yet, significant waning of specific immunity within six months after 2nd vaccination, along with higher incidence of breakthrough infections associated with the time elapsed since 2nd vaccination raises concerns regarding the durability of immunity also after 3rd vaccination. We assessed virus-specific serum antibody and T cell response in the blood after vaccination with the mRNA vaccine BNT162b2 in more than 50 individuals older than 80 years. All old adults demonstrated a strong humoral response to 3rd vaccination which was at average higher and waned slower than the response to 2nd vaccination, indicative of enhanced humoral immunity. In contrast, their respective T cell response quantitatively limited to the level obtained after 2nd vaccination, with similar waning over time and no evidence for enhanced IFNg production. Because BNT162b2-mediated protection from the Omicron variant relies more on T cells than antibodies, our findings raise concern on the durability of protection from the Omicron variant by BNT162b2 in the senior population.

## Main text

A third mRNA-based “booster” vaccination is the currently favored strategy to maintain protection against SARS-CoV-2 infection. Yet, significant waning of specific immunity within six months after 2^nd^ vaccination ^1^, along with higher incidence of breakthrough infections associated with the time elapsed since 2^nd^ vaccination ^2,3^ raises concerns regarding the durability of immunity also after 3^rd^ vaccination.

We compared the specific humoral and cellular response (Fig.1A-C) after 3^rd^ vs. 2^nd^ vaccination with BNT162b2 in a cohort of adults aged >80 years (sTable1,2 ^4^) at risk for severe COVID-19 and immune senescence. Our data demonstrate the induction of marginally higher spike S1-specific blood IgG concentrations two weeks after 3^rd^ than after 2^nd^ vaccination (sFig.1A, Fig.1D). By contrast, functionally relevant RBD-specific IgG (Fig.1B) and SARS-CoV-2-neutralizing antibody (sFig.1B) titers were substantially increased (Fig.1D) after 3^rd^ compared to 2^nd^ vaccination, reflecting enhanced antibody production and/or affinity maturation.

**Figure 1.**
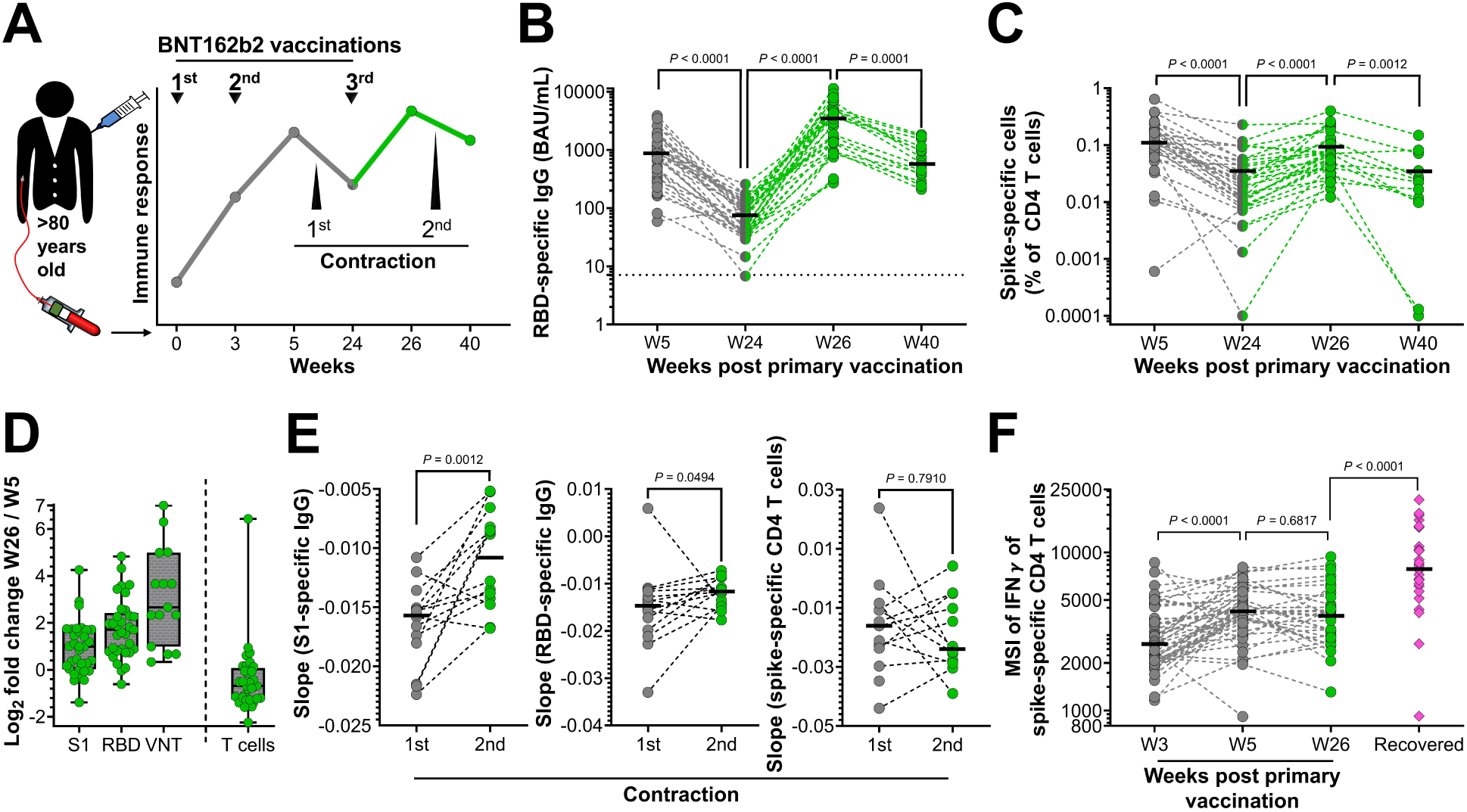
Humoral and cellular SARS-CoV-2 immunity in >80 year-old donors after a third “booster” BNT162b2 vaccination. Study overview (A). Immune response kinetics were followed in older adults (sTable 1 and 2) in the course of vaccinations with BNT162b2. Green color indicates data related to the third dose of BNT162b2. Vaccinations (arrows) and blood samplings (dots) are indicated. SARS-CoV-2 RBD-specific serum IgG levels (B), frequencies of SARS-CoV-2 spike-specific CD4 T cells identified as CD40L+IFNγ+ CD4 T cells after overnight stimulation of PBMC with SARS-CoV-2 spike peptides (C) were analyzed. For W5, W24, W26, W40: *n* = 35, 36, 34, 15 (B) and *n* = 34, 35, 33, 13 (C), respectively. Log2 of the fold change in peak response values for spike (S1 subunit)- and RBD-specific IgG (*n* = 33), neutralizating antibody titers (*n* = 15), and spike-specific T cell frequencies (*n* = 32) in participants after 2^nd^ and 3^rd^ vaccination (D). Slope of the declining peak response (sFig.2) after 2^nd^ (1^st^ contraction) or 3^rd^ (2^nd^ contraction) vaccination, calculated for S1- and RBD-specific IgG titers (*n* = 14) and for spike-specific T cell frequencies (*n* = 12) (E). Comparison of the median staining intensity (MSI) of IFNγ of spike-specific CD4 T cells of vaccinated participants determined at W3 (*n* = 51), W5 (*n* = 51), and W26 (*n* = 33), and of >80 year old donors recovered from COVID-19 (not vaccinated, *n* = 30, magenta) (F). Control stimulations of PBMC with staphylococcal enterotoxin B (sFig.3) confirmed the stability of the T cell stimulation conditions over time. Each symbol represents data of one donor at one time point. Horizontal lines indicate median values of data points in each column. The dotted horizontal line (B) indicates the cutoff for antibody positivity at 7.1 BAU / mL. *P* values were determined by two-tailed Wilcoxon matched-pairs signed rank test throughout except when comparing vaccinated (W26) vs recovered groups (F, two-tailed Mann-Whitney test). VNT, virus neutralization titer; RBD, receptor binding domain; IFN, interferon; BAU, binding antibody units.

In contrast, spike-specific CD4 T cell frequencies reached similar levels after 2^nd^ and 3^rd^ vaccination (Fig.1C,D). After the respective acute response, frequencies returned to approximately pre-vaccination levels with similar rates of decline after 2^nd^ and 3^rd^ vaccination (week 24 and 40, respectively, Fig.1C,E, sFig.2). Quantified cytoplasmic expression of the effector cytokine IFNγ indicated functional enhancement of spike-specific T cells upon 2^nd^ but not further upon 3^rd^ vaccination, while more cytoplasmic IFNγ was found in spike-specific CD4 T cells from senior adults recovered from COVID-19 (Fig.1F). Thus, even a third BNT162b2 dose failed to induce durably enhanced quantities of spike-specific T cells and a functional quality reached after natural infection.

Concentrations of S1- and RBD-specific IgG also declined from the acute responses at week 5 and week 26, but clearly at a lower rate after 3^rd^ (week 40) compared to 2^nd^ (week 24) vaccination (Fig.1B,E, sFig.1A, sFig.2), yielding persistently enhanced IgG quantity and/or quality after the 3^rd^ vaccination. We conclude that a third vaccination against SARS-CoV-2 in older adults, while establishing immunity in primary non-responders^4^, induces a durably escalated humoral response in the bulk of vaccinees for at least three months, indicating longer lasting humoral immunity. However, BNT162b2-induced immune reactivity against the emerging Omicron variant is predominantly T cell-mediated ^5,6^. Thus, the apparent inability of older adults to develop and maintain high levels of fully matured SARS-CoV-2-reactive T cells after 3^rd^ vaccination is of concern. Additional immunizations with vaccines designed to target “variants of concern” and improve T cell reactivity will be required to satisfactorily protect senior adults from COVID-19.

## Data Availability

All data produced in the present study are available upon reasonable request to the authors

## Supplementary material

**sTable 1.** Summary of vaccinated donors characteristics.

| Group | Total donors | Sex | Age range | Age mean / SD |
| --- | --- | --- | --- | --- |
| **W0, W3** | *n* = 52 | female: 31 (60%)<br>male: 21 (40%) | 80 – 97 years | 84.0 / 3.7 |
| **W5** | *n* = 49 | female: 30 (61%)<br>male: 19 (39%) | 80 – 97 years | 84.0 / 3.7 |
| **W24** | *n* = 37 | female: 23 (62%)<br>male: 14 (38%) | 80 – 97 years | 83.4 / 3.7 |
| **W26** | *n* = 34 | female: 22 (65%)<br>male: 12 (35%) | 80 – 97 years | 83.4 / 3.6 |
| **W40** | *n* = 15 | female: 11 (73%)<br>male: 4 (27%) | 80 – 97 years | 84.7 / 4.6 |

**sTable 2.**
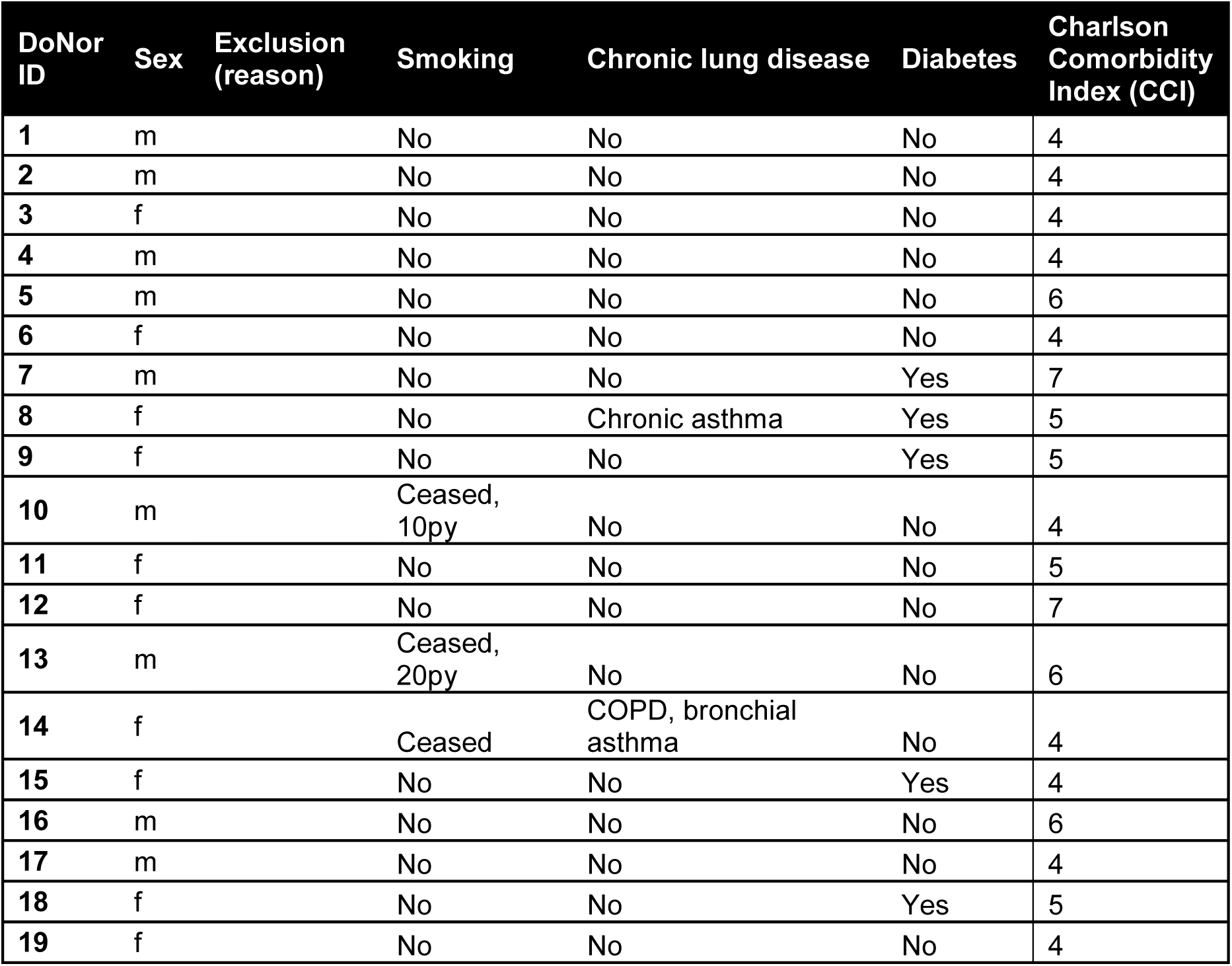
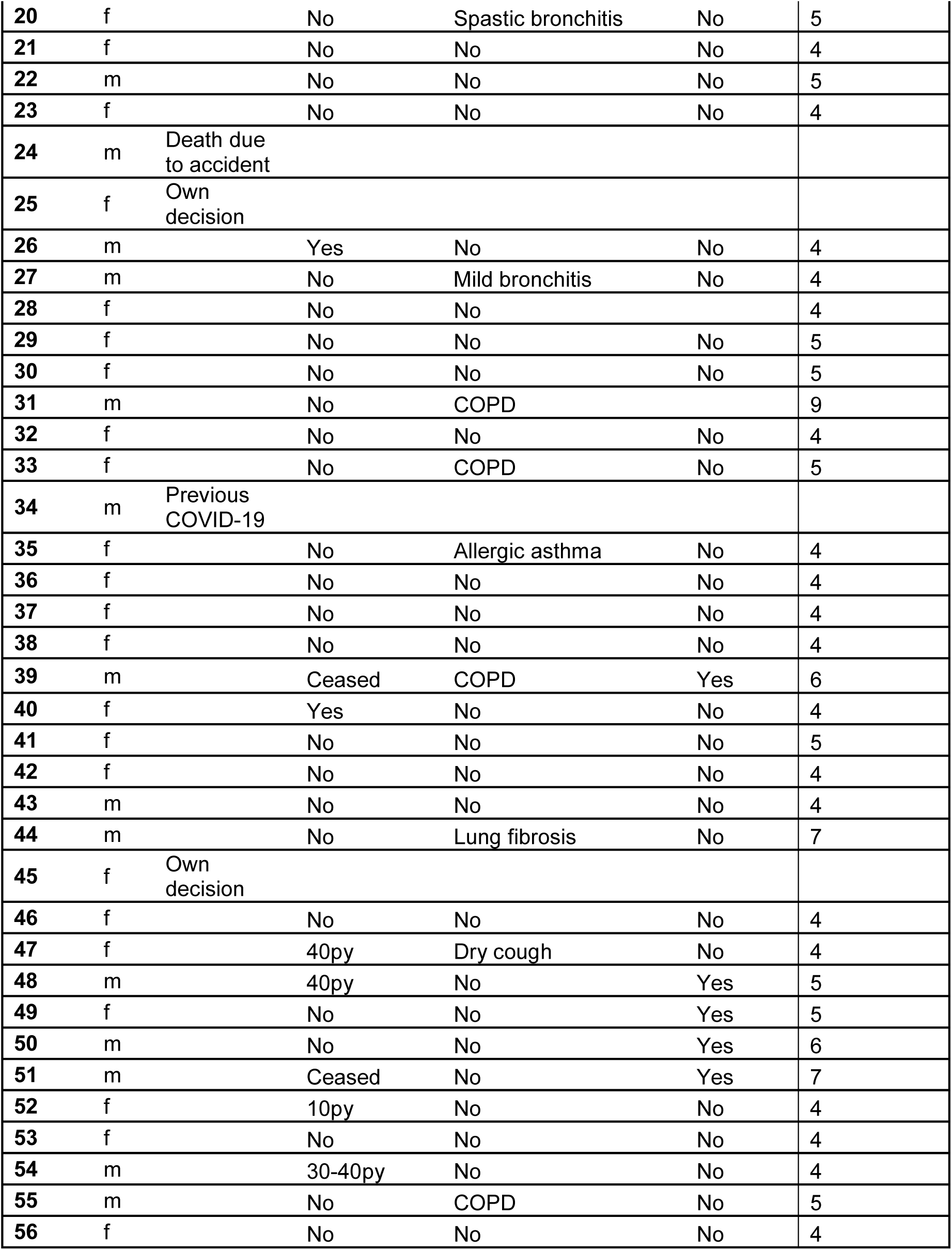
Baseline characteristics of vaccinated participants.

**sFigure 1.**
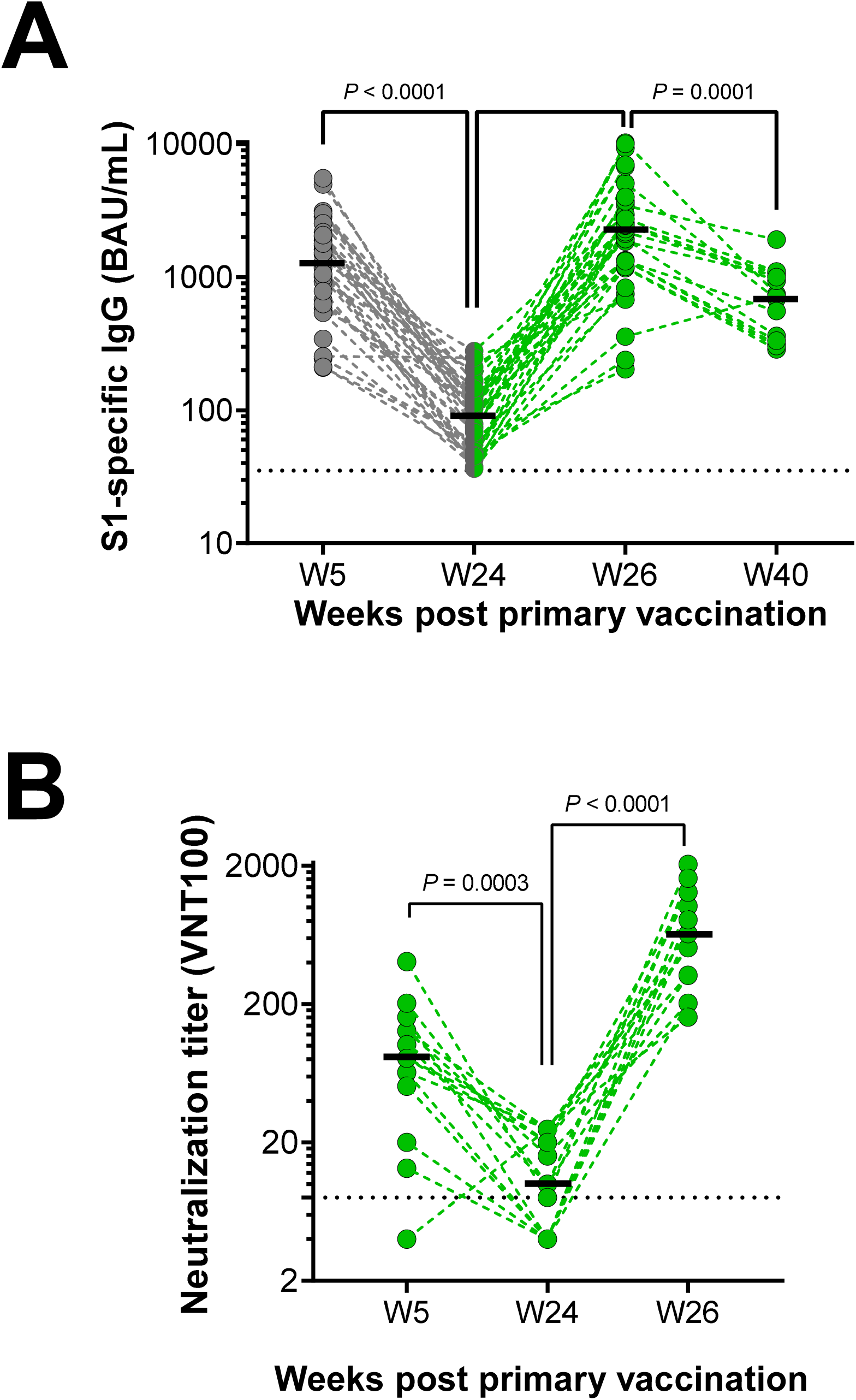
Increased antibody response in >80 year-old donors after receiving a third dose of the BNT162b2 vaccine. SARS-CoV-2 spike (S1-sbunit)-specific serum IgG levels (A) and serum titers of 100% virus neutralization for SARS-CoV-2 wild type B.1 (VNT 100) (B) were analyzed in donors aged >80 years at the indicated time points after primary vaccination with BNT162b2 (see Figure 1A for study overview). Each symbol represents one donor. Horizontal lines indicate medians, horizontal dotted lines indicate the cutoff for antibody positivity at 35.2 BAU / mL (A) and the lower detection limit for the VNT 100 assay at a reciprocal titer of 8 (B). *n* = 35, 36, 34, 15 donors were analyzed on W5, W24, W26, W40, respectively in (A); *n* = 15 donors on all time points in (B). *P* values were determined using the two-tailed Wilcoxon matched-pairs signed rank test.

**sFigure 2.**
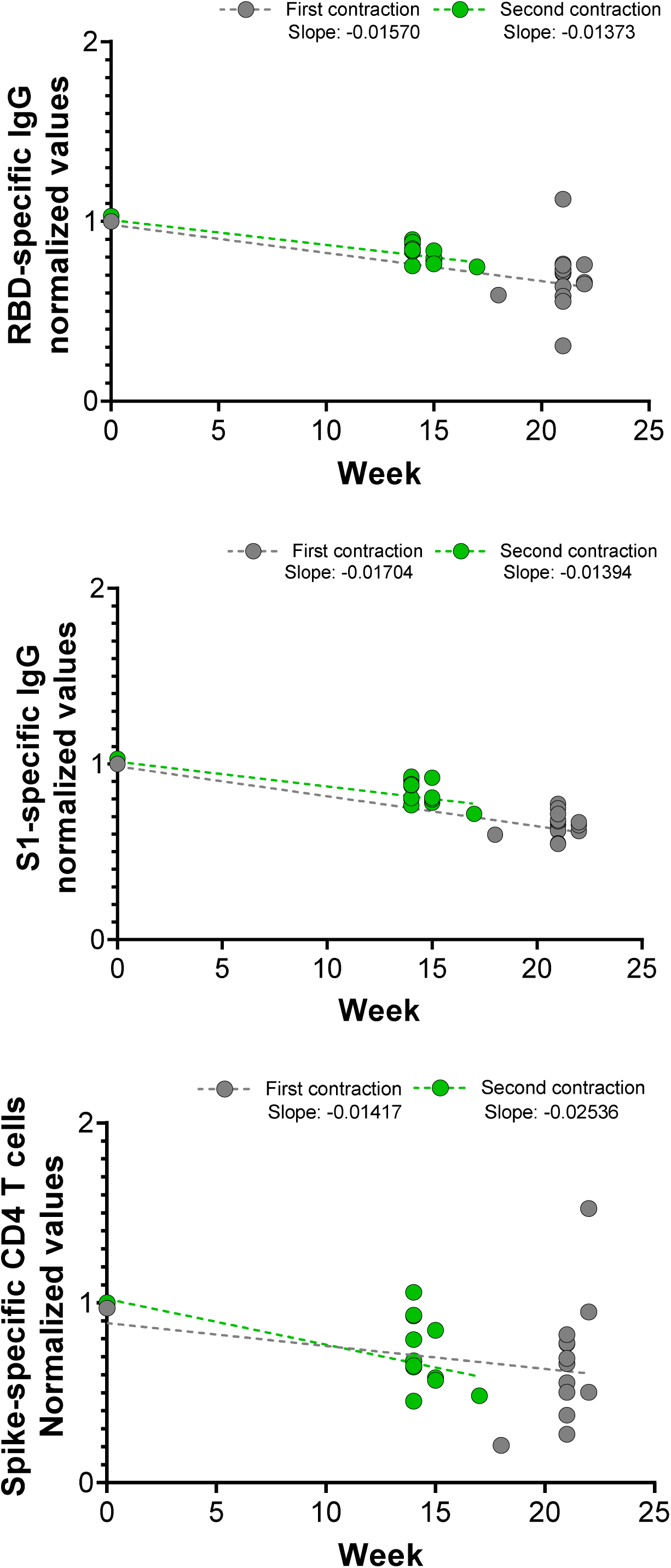
Contraction of SARS-CoV-2 specific CD4 T cell and humoral immunity after second and third “booster” vaccination with BNT162b2. Normalized slopes of serum SARS-CoV-2 RBD-specific IgG, S1-specific IgG, and circulating SARS-CoV-2 spike-specific CD4 T cells, after the second dose (first contraction, grey dots) and after the third dose (second contraction, green dots) of BNT162b2. Weeks indicated on the x axis are weeks after respective peak responses. Fitted linear regression lines are shown as dashed lines. Slopes were calculated by linear regression from log-transformed data. Data for each donor and each contraction was normalized to a value of 1 at week 5 (first contraction) and week 26 (second contraction). Slopes were calculated from 14 donors (RBD- and S1-specific IgG serum responses) and 12 donors (spike-specific CD4 T cell responses.

**sFigure 3.**
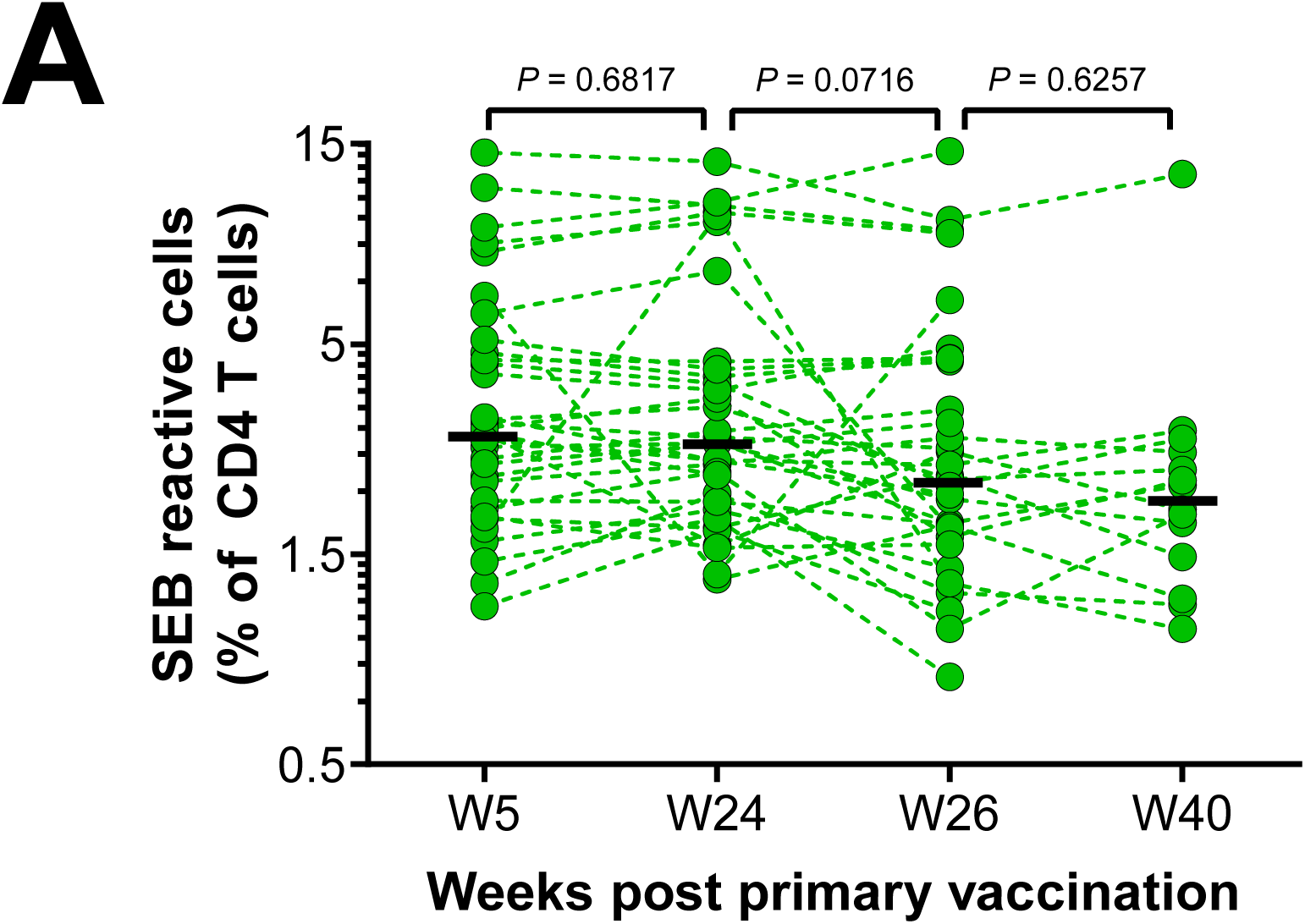
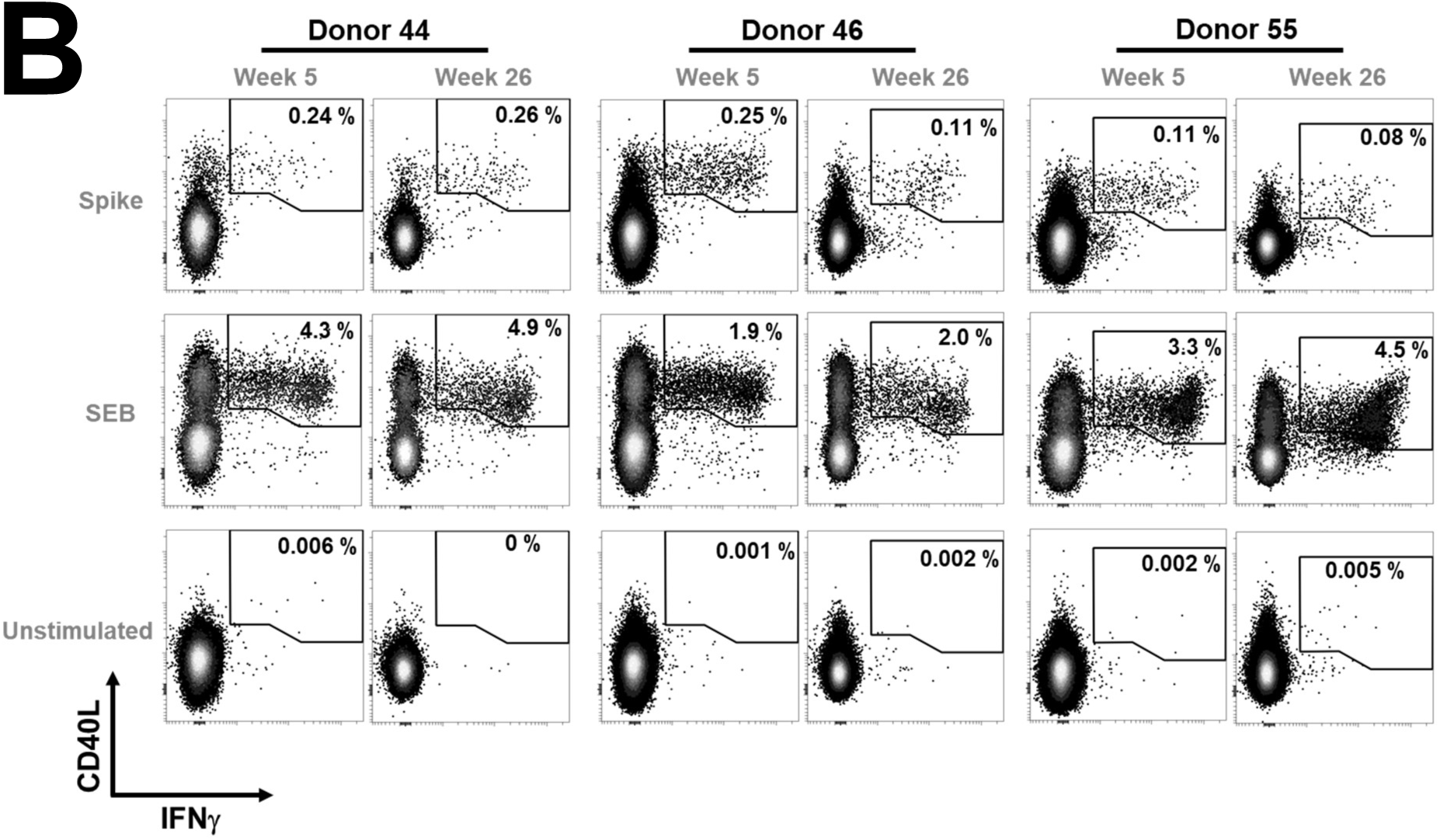
Blood CD4 T cell response to SEB in >80 year-old vaccinated donors and examples of flow cytometric analysis. Blood was collected at the indicated time points post primary vaccination. Vaccinations with BNT162b2 were performed at week 0, week 3 and week 24 (Fig. 1A). Frequencies of CD40L+IFNγ+ CD4 T cells after culture of PBMC in the presence of the superantigen staphylococcal enterotoxin B (SEB) are shown (A). *n* = 34, 35, 34, 14 donors were analyzed on W5, W24, W26, W40, respectively. *P* values were determined by two-tailed Wilcoxon matched-pairs signed rank test. Representative analysis of flow cytrometry data depicting the detection of CD40L+IFNγ+ CD4 T cells in three donors at week 5 and week 26 are shown in (B). PBMC were stimulated in the presence or absence of spike protein peptide pool or SEB. Data of CD4 T cells are shown; frequencies of gated cells are indicated.

## Material and Methods

### Study participants

Up to six blood samples were obtained from participants aged >80 years by venipuncture as shown in Fig. 1A. Samples were collected before first (week 0, W0), and three weeks (W3, right before 2^nd^ vaccination), five weeks (W5), average 24 weeks (W24, before 3^rd^ vaccination), average 26 weeks (W26) and 40 weeks (W40) after first vaccination by injection of Tozinameran (BNT162b2 vaccine, Comirnaty®) in the deltoid muscle, at a vaccination center in Germany (sTables 1 and 2).

Analyses of vaccinated donors were performed between March 2021 and January 2022. In May 2021, blood samples were obtained by venipuncture from unvaccinated donors >80 years of age living in a retirement home, having recovered from COVID-19 after an outbreak with SARS-CoV-2 variant B.1.221 in January 2021. All donor provided informed consent to participate in the study. Few participants were later excluded from the study for reasons unrelated to the study (sTable 2). Additionally, data of one donor was excluded from the whole T cell analysis due to strong erythrolysis (Polycythemia Vera) and data of one donor excluded on W40 due to sample clogging during the T cell assay. The study of patients with COVID-19 and vaccinations against COVID-19 was approved by the ethics committee of the medical faculty of the Philipps-University Marburg (study number 40/21-12032021).

### Sample processing and clinical lab

Blood serum was isolated from Serum Separator Clot Activator tubes (Greiner Bio-One GmbH, Frickenhausen, Germany) according to the manufacturer’s instructions, and stored frozen until analysis.

Peripheral blood mononuclear cells (PBMC) were isolated from fresh heparinized whole blood by density gradient centrifugation over Pancoll human (Pan Biotech, Aidenbach, Germany) after dilution with an equal volume of PBS at room temperature. PBMC were washed twice (500 x g, 10 min, 4 °C) in cold PBS supplemented with 0.2% BSA, counted manually, and resuspended in RPMI 1640 media (Gibco, Life Technologies, Carlsbad, CA) supplemented with penicillin, streptomycin, and 10% human AB serum (all Sigma, St. Louis, MO) at 5 × 10^6^ cells / mL.

### Assessment of antigen-specific T cells (ART)

Antigen-reactive T cell responses were analyzed using a protocol based on previous work^1^. 500 µL media containing 5 × 10^6^ PBMC were transferred into 12 mL round-bottom tubes (Greiner Bio-One GmbH, Frickenhausen, Germany) and stimulated with either SARS-CoV-2 spike protein peptide mix (wildtype, Miltenyi Biotec), SEB (0.7 µg / mL, kindly provided by Prof. Bernhard Fleischer, Bernhard Nocht Institute of Tropical Medicine, Hamburg, Germany), or with an equal volume of water as a control, in the presence of anti-CD28 (5 µg / mL) and monensin (1 µg / mL) for 12 hours under humid conditions in a 5 % CO2 atmosphere. Brefeldin A (1 µg / mL) was added 2 hours after the start of the stimulation. The stimulation was stopped by adding 2 nM EDTA. PBMC were harvested, transferred to a new 15 mL centrifuge tube, washed with 10 mL PBS / 0.2 % BSA, and pelleted for 10 min at 490 x g, at 4 °C. Dead cell labeling was performed by resuspending the cell pellet in 500 µL PBS supplemented with 1:1000 amine-reactive Zombie Aqua™ Fixable Viability dye (Biolegend), incubated for 20 min in the dark at room temperature. PBS / 0.2 % BSA was added to quench the remaining reactive dye. After washing with 2 mL PBS / 0.2 % BSA, and pelleting for 10 min, at 490 x g, at 4 °C, PBMC were fixed for 20 minutes using 2% formaldehyde solution (Thermo Scientific, Germany) in the dark and washed twice with 2 mL PBS / 0.2 % BSA (10 min, 700 x g, 4 °C). Cell pellets were resuspended in 200 µL PBS / 0.2 % BSA, transferred into a V-bottom-96 well plate (Sarstedt), and centrifuged (700 x g, 5 min, 4 °C). After discarding the supernatants, pellets were resuspended in 50 µL Brilliant Violet Staining Buffer (Biolegend) supplemented with antibodies including anti-CD4-PECy7 (1:400 dilution) for the detection of CD4 T cells, anti-CD8-BV570 (1:100 dilution), and anti-CD19/123/33-BV510 (all from Biolegend at 1:50,1:50, 1:400 dilution, respectively) for the exclusion of CD8 T cells, monocytes and other myeloid cells, B cells, basophils, and plasmacytoid dendritic cells, and incubated 30 minutes in the dark, at room temperature. Afterwards, cells were washed once with 200 µL PBS / 0.2% BSA, and centrifuged (700 x g, 5 min, 4 °C). Cell pellets were then washed twice in 200 µL permeabilization buffer (diluted using Millipore water from 10 x concentrated stock buffer, ThermoFisher, Waltham, MA), and finally resuspended in 50 µL permeabilization buffer supplemented with antibodies targeting CD40L (1:100 dilution conjugated to BV421, Biolegend), and intracellular molecules including anti-IFNγ-R718 (1:100 dilution BD Biosciences, San Jose, CA), and incubated for 30 minutes at room temperature in the dark. Then, cells were washed once in 200 µL permeabilization buffer, once in PBS / 0.2 % BSA, and stored at 4 °C in PBS / 0.2 % BSA until acquisition on a MACSQuant 16 flow cytometer (Miltenyi Biotec, Bergisch Gladbach, Germany).

FlowJo version 10 (BD, Ashland, OR) and OMIQ.ai (Santa Clara, CA) were used for analyzing flow cytometry data. Flow cytometry standard (FCS) files underwent quality control and, where applicable, anomaly removal by FlowAI ^2^, and data of live CD4+ T lymphocytes were gated according to FSC and SSC parameters and their CD4^+^CD8^-^CD19^-^CD123^-^CD33^-^LD-Aqua^low/-^ phenotype. Cell aggregates were removed by gating according to FSC-H/FSC-A and SSC-H/SSC-A parameters. Antigen-reactive CD4 T cells (ART) were defined as CD40L+ IFNγ+ expressing cells after spike peptide stimulation, and their frequencies were determined among total CD4 T cells.

For background correction, frequencies of CD40L+IFNγ+ T cells among CD4 T cells from unstimulated control samples were subtracted from ART frequencies for each individual donor. Whenever this difference was equal to or below 0%, frequencies were set to 0.0001%, a value exceeded by all measurements in which ART were detectable, in order to allow the display of data on a logarithmic scale in Figure 1.

### Quantification of SARS-CoV-2-specific antibodies

Serum IgG antibodies against the receptor-binding domain (RBD) of the SARS-CoV-2 spike protein were quantified using the automated SARS-CoV-2-IgG-II-Quant-Assay on the Abbott Alinity i analyzer (Abbott, Wiesbaden, Germany), following the manufacturer’s protocol. Results obtained in arbitrary units / mL (AU / mL) were converted into binding antibody units (BAU) / mL by multiplication with the factor 0.142, according to the manufacturer’s instructions. Results in BAU / mL are calibrated against the “First WHO International Standard for anti-SARS-CoV-2 immunoglobulin (NIBSC code: 20/136)”. The lower cutoff for this assay is at 7.1 BAU / mL. Sera exceeding the detection range of the assay (40,000 AU /mL) were automatically pre-diluted 1:2 by the device and measured again.

Serum IgG directed against the S1 subunit of the SARS-CoV-2 spike protein was quantified in BAU / mL using the Anti-SARS-CoV-2-QuantiVac ELISA (Euroimmun, Lübeck, Germany). Pipetting was performed on the automated EuroLab Workstation (Euroimmun), following the manufacturer’s protocol. Primary data obtained in relative units per mL were converted into binding antibody units (BAU) / mL by multiplication with the factor 3.2, according to the manufacturer’s instructions. Results in BAU / mL are calibrated against the “First WHO International Standard for anti-SARS-CoV-2 immunoglobulin (NIBSC code: 20/136)”. The lower cutoff for this assay is at 35.2 BAU / mL. Sera exceeding the detection range of the assay were manually pre-diluted 1:10 and/or 1:50 and measured again.

### SARS-CoV-2 neutralization tests (VNT-100)

Human sera were heat-inactivated for 30 min at 56 °C and diluted in a two-fold dilution series in 96-well cell culture plates (1:4 to 1:512 for week 24 sera, 1:8 to 1:1024 for week 26 sera). One hundred plaque-forming units (PFU) of SARS-CoV-2 were added in the same volume to the serum dilutions. The German SARS-CoV-2 virus isolate BavPat1/2020 (European Virus Archive Global #026 V-03883, Genbank: MZ558051.1) was used. The sequence of the viruses was confirmed. Following incubation at 37 °C for 1 h, approximately 20,000 Vero C1008 cells (ATCC, Cat. no. CRL-1586, RRID: CVCL_0574) were added. Plates were then incubated at 37 °C in a 5 % CO2 atmosphere, and cytopathic effects were evaluated at day 4 post infection. Neutralization was defined as the absence of cytopathic effects in the serum dilutions. The reciprocal neutralization titer was calculated from the highest serum dilution without cytopathic effects as a geometric mean based on three replicates. The lower detection limit of the assay is 8 (reciprocal titer) for week 24 sera and 16 for week 26 sera, corresponding to the first dilution of the respective serum. Two positive controls were used as inter-assay neutralization standards and quality control for each test. Neutralization assays were performed in the BSL-4 laboratory of the Institute of Virology at Philipps University Marburg, Germany.

### Statistical analysis details

Prism version 9 (GraphPad software, San Diego, CA) was used to display data, and perform descriptive statistics and significance testing. To determine differences between cohorts in Fig. 1F (vaccinated group W26 vs recovered group), a two-tailed Mann–Whitney test was applied to calculate the P value. For assessing donor-specific responses over time, the two-tailed Wilcoxon matched-pairs signed-rank test was used. Slopes reflecting immune contractions were calculated by linear regression from log-transformed data. Data for each donor and each contraction was normalized to 1 for week 5 (first contraction) and week 26 (second contraction). Donors selected to evaluate contractions (slopes) were those who participated also in W40. Slopes were calculated from 14 donors (RBD and spike IgG serum responses) and 12 donors (spike-specific CD4 T cell responses).

## Acknowledgements

The authors wish to thank all participating donors and especially the vaccination center in Germany, and its Chief Manager K. Oerder for excellent support throughout this study.

